# Cost comparison of unplanned hospital admissions from care home and community settings: A retrospective cohort study using routinely collected linked data

**DOI:** 10.1101/2024.06.27.24309582

**Authors:** C Geue, G Ciminata, G Reddy Mukka, D Mackay, J Lewsey, JM Friday, R Dundas, QB Tran, D Brown, F Ho, C Hastie, M Fleming, A Stevenson, C du Toit, S Padmanabhan, JP Pell

**Affiliations:** University of Glasgow

## Abstract

People living in care homes often have multiple morbidities and complex healthcare needs, potentially leading to more frequent healthcare utilisation (planned and unplanned) and increased costs. Unscheduled hospital attendance from a care home setting is often regarded as system failure, futile and inefficient in terms of resource use. However, there is a lack of evidence on the costs associated with these attendances. This retrospective cohort study aims to estimate these costs and provides a comparison by usual place of residence. Data were obtained from NHS Greater Glasgow and Clyde Safe Haven reference datasets. Individual-level record linkage between Trak ED, recording emergency admissions, and other routine healthcare datasets was carried out. Healthcare costs were estimated using a two-part model. The first part used a probit model to estimate the probability of positive healthcare resource utilisation, and the second part used a GLM to estimate costs, conditional on costs being positive. Annual mean costs were higher for care home residents than community-dwellers overall and in both men and women and all deprivation quintiles. No significant difference in costs was observed for care home residents who were younger than 65 years and those with no comorbidity. Our results indicate a notable increase in healthcare expenditure for individuals residing in care homes compared to those living in the community following unplanned acute care incidents, emphasising the importance of developing interventions that are specifically designed to meet the unique requirements of this demographic.

## INTRODUCTION

About half a million people in the UK reside in care homes, with the majority being older adults [1;2]. In Scotland, about 40,000 people live in care homes long-term [3]. Care home residents often have multiple morbidities and complex healthcare needs, leading to more frequent hospitalisations (planned and unplanned), extended hospital stays, and increased hospital costs [4;5]. Frequent emergency department (ED) visits from a care home setting have been reported, with an average of 1.8 visits per care home resident per year, with only about half of these visits leading to hospital admission [6]. Frequent use of NHS resources by care or nursing home residents was also reported in a small prospective study from the UK, which did report resource use, but did not provide cost estimates [7].

Unscheduled hospital attendance from a care home setting is often regarded as system failure, futile and inefficient in terms of resource use [8-10]. However, there is also recognition that transfer decisions are complex, multi-faceted and not solely related to clinical needs [11]. A recent systematic review summarised evidence in terms of effective interventions to reduce unplanned hospital attendances from a care home setting, but also highlighted the lack of economic evidence on the implementation of such interventions [12]. In addition to a lack of evidence in terms of resource use and associated costs, there is also uncertainty as to whether people attending an ED from a care home setting differ in terms of outcomes from people attending from a community setting. Certain risk factors, including male sex, age and presence of certain comorbidities have been identified as predictors of transfer to ED from a care home setting [13]. These predictors can usefully inform targeted interventions to reduce transfers, however, a detailed analysis of cost implications is needed in addition to health and care related outcome measures if interventions are to be evaluated in terms of their cost-effectiveness. Evidence from the US shows that the cost of potentially avoidable hospitalisations for long-stay care home residents was estimated to be $4,206 higher per case than for those managed in the care home [14].

This retrospective cohort study aims to estimate the costs associated with unscheduled hospital attendance and provide a comparison by usual place of residence (care home vs community). Our analysis will also help to assess current data capabilities and thus inform future data initiatives to better understand patient pathways, in particular for long-term care home residents.

## METHODS

### Study Design

We conducted a retrospective cohort study of ED attendees, comparing those who resided in care homes with a community-dwelling comparison group matched 1:1 by age, sex, date of attendance of index ED visit, and specialty.

### Data Sources

Data were obtained from NHS Greater Glasgow and Clyde (NHS GGC) Safe Haven reference datasets from July 2017 to July 2021. Trakcare is an electronic patient record system on unscheduled care and emergency healthcare services [15] and includes Trak ED, an ED database, which was used to identify ED visits. The Community Health Index, a unique identifier, was used to perform individual-level record linkage between Trak ED and four other databases prior to anonymisation: i) the Prescribing Information System (PIS), ii) demographic data, iii) National Records for Scotland (NRS) mortality records, and iv) Scottish Morbidity Records 01 (SMR01) which cover hospital admissions. Externally available lookup files with unit costs were checked by the NHS GGC data provider and uploaded for analysis. The PIS database included a care home flag and was used to ascertain exposure status [16]. The demographic database was used to ascertain the confounders used for matching and adjustment. The SMR01 database provided information on length of hospital stay. The Charlson Comorbidity Index is provided using information on relevant ICD-10 codes from across different routinely held health records in NHS GGC.

### Outcome Measure

Annual cost profiles for care home residents and people attending ED from the community were developed, including all subsequent healthcare resource use following the index ED visit (inpatient stays, repeat ED attendance). Published per diem unit costs were used to value respective resource use items [17]. These were mapped according to specialty and multiplied by length of stay in hospital. Costs associated with ED attendance were also taken from published unit costs [17].

### Statistical Analyses

Participant characteristics were summarised using frequencies and percentages. Healthcare costs were estimated using a Generalised Linear Model (GLM).

Regression models were adjusted for age, sex, an indicator of area socioeconomic status (Scottish Index of Multiple Deprivation (SIMD 2016) represented as quintiles with one denoting the most deprived areas and five denoting the least deprived areas, respectively), and the number of co-morbidities. A two-part model was used to model costs, accounting for the skewed distribution of costs data. The first part used a probit model to estimate the probability of positive healthcare resource utilisation, and the second part used a GLM with gamma distribution and log link to estimate costs, conditional on costs being positive. Both modelling parts included identical explanatory variables. Separate models were run for each cost component (inpatient care and ED attendance), with a further model estimating overall costs. Effect sizes are presented as cost-ratio regression coefficients with their respective 95% CI. We used recycled predictions to estimate and compare marginal effects for each of the included covariates, holding other covariates at their mean values.

All statistical analyses were undertaken using Stata MP, version 18 (StataCorp).

## Ethics & Data Extraction Methods

Delegated research ethics approval was granted for linkage to National Health Service (NHS) patient data by the Local Privacy and Advisory Committee at NHS Greater Glasgow and Clyde. Cohorts and de-identified linked data were prepared by the West of Scotland Safe Haven Research Database at NHS Greater Glasgow and Clyde (IRAS Project ID 321198, REC reference 22/WS/1063).

## RESULTS

The study cohort comprised 11,568 individuals with 5,784 each accessing ED from a care home setting or a community setting.

A substantial proportion, 30.8% of care home residents and 36.6% of community residents, had five or more ED visits during the observational period (2017-2021) (Table 1). We observed a significantly higher proportion of people with more than one comorbidity (60.3%) attending ED from a care home setting, compared to people with more than one comorbidity attending from a community setting (53.4%).

**Table 1:** Cohort Characteristics.

|  | Care home resident |  | Community-dweller |  |  |
| --- | --- | --- | --- | --- | --- |
|  | N | Percentage | N | Percentage |  |
| **Sex** |  |  |  |  | P=0.517 |
| Female | 3,523 | 60.91 | 3,557 | 61.50 |  |
| Male | 2,261 | 39.09 | 2,227 | 38.50 |  |
| **Age Category (years)** |  |  |  |  | P<0.001 |
| ≤50 | 478 | 8.26 | 409 | 7.07 |  |
| 51 – 55 | 119 | 2.06 | 167 | 2.89 |  |
| 56 – 60 | 167 | 2.89 | 148 | 2.56 |  |
| 61 – 65 | 172 | 2.97 | 227 | 3.92 |  |
| 66 – 70 | 306 | 5.29 | 297 | 5.13 |  |
| 71 – 75 | 475 | 8.21 | 532 | 9.20 |  |
| 76 – 80 | 785 | 13.57 | 820 | 14.18 |  |
| 81 – 85 | 1,108 | 19.16 | 1,173 | 20.28 |  |
| 86 – 90 | 1,204 | 20.82 | 1,159 | 20.04 |  |
| ≥91 | 970 | 16.77 | 852 | 14.73 |  |
| **Deprivation Quintile (SIMD)** |  |  |  |  | P<0.001 |
| 1 – most deprived | 2,502 | 43.26 | 2,618 | 45.26 |  |
| 2 | 733 | 12.67 | 1,061 | 18.34 |  |
| 3 | 1,086 | 18.78 | 761 | 13.16 |  |
| 4 | 842 | 14.56 | 641 | 11.08 |  |
| 5 – least deprived | 621 | 10.74 | 703 | 12.15 |  |
| **Number of ED visits** |  |  |  |  |  |
| 1 | 1,193 | 20.63 | 963 | 16.65 |  |
| 2 | 1,173 | 20.28 | 1,001 | 17.31 |  |
| 3 | 946 | 16.36 | 911 | 15.75 |  |
| 4 | 670 | 11.58 | 737 | 12.74 |  |
| ≥5 | 1,781 | 30.79 | 2,115 | 36.57 |  |
| **Comorbidities** |  |  |  |  | P<0.001 |
| 0 | 746 | 12.90 | 1,480 | 25.59 |  |
| 1 | 1,548 | 26.76 | 1,216 | 21.02 |  |
| More than 1 | 3,490 | 60.34 | 3,088 | 53.39 |  |
| **ED re-attendance within 30 days of ED discharge** |  |  |  |  | p=0.333 |
| No | 2,757 | 47.67 | 2,705 | 46.67 |  |
| Yes | 3,027 | 52.33 | 3,079 | 53.23 |  |

In terms of re-attending ED within 30 days of being discharged from ED, no significant difference was found between groups.

Annual estimated costs per patient for attending ED as well as costs related to any hospitalisation that occurred following the incident ED visit are presented in Table 2 (full regression output for both modelling parts can be found in supplementary table S1).

**Table 2:** Annual total cost estimates per patient: overall and marginal by covariate using recycled predictions.

|  | **Community-dweller**<br>**£ (95% CI)** | **Care home resident**<br>**£ (95% CI)** |
| --- | --- | --- |
| **Overall cost** |  |  |
| Unadjusted | 4,263 (4,119 to 4,406) | 4,991 (4,796 to 5,186) |
| Adjusted | 4,786 (4,631 to 4,941) | 4,345 (4,177 to 4,512) |
| **Sex** |  |  |
| Female | 4,082 (3,935 to 4,230) | 5,303 (5,063 to 5,544) |
| Male | 4,077 (3,900 to 4,254) | 5,296 (5,050 to 5,541) |
| **Age Category (years)** |  |  |
| ≤50 | 8,190 (6,555 to 9,825) | 11,807 (8,899 to 14,715) |
| 51-55 | 6,946 (4,796 to 9,095) | 10,051 (6,275 to 13,828) |
| 56-60 | 6,056 (4,772 to 7,339) | 8,166 (5,992 to 10,340) |
| 61-65 | 4,631 (3,977 to 5,286) | 6,077 (5,001 to 7,154) |
| 66-70 | 4,597 (4,106 to 5,089) | 5,784 (5,104 to 6,465) |
| 71-75 | 4,087 (3,768 to 4,407) | 5,356 (4,869 to 5,843) |
| 76-80 | 4,094 (3,830 to 4,358) | 5,197 (4,790 to 5,604) |
| 81-85 | 4,037 (3,803 to 4,271) | 4,659 (4,386 to 4,932) |
| 86-90 | 4,170 (3,936 to 4,403) | 4,932 (4,639 to 5,224) |
| >90 | 4,171 (3,813 to 4,529) | 4,654 (4,305 to 5,004) |
| **Comorbidities** |  |  |
| 0 | 1,809 (1,669 to 1,949) | 2,073 (1,892 to 2,255) |
| 1 | 3,376 (3,185 to 3,567) | 3,923 (3,676 to 4,169) |
| More than 1 | 5,989 (5,739 to 6,238) | 7,038 (6,724 to 7,352) |
| **Deprivation Quintile (SIMD)** |  |  |
| 1 – most deprived | 4,181 (4,014 to 4,348) | 5,445 (5,197 to 5,694) |
| 2 | 4,358 (4,085 to 4,630) | 5,677 (5,284 to 6,070) |
| 3 | 3,883 (3,632 to 4,133) | 5,067 (4,732 to 5,402) |
| 4 | 4,067 (3,775 to 4,359) | 5,307 (4,905 to 5,709) |
| 5 – least deprived | 3,458 (3,197 to 3,719) | 4,517 (4,159 to 4,874) |

The annual average costs were higher among care home residents than community-dwellers overall and in both men and women and all deprivation quintiles. Whilst the same pattern was observed in people over 65 years of age and with at least one comorbidity, there was no significant difference in costs for care home residents who were younger than 65 years of age and those with no comorbidity.

Annual cost estimates separated by cost component (inpatient and ED) can be found in supplementary tables 2 and 3.

## DISCUSSION

This is the first study to utilise comprehensive, linked electronic health records to compare the economic cost of unplanned ED attendances among care home residents with community-dwellers. Our analysis provides useful evidence for decision making on the differences in healthcare costs incurred, depending on people’s usual place of residence. Future economic evaluations of interventions that aim to reduce the number of ED attendances will be able to utilise these estimates in their evaluations.

Healthcare costs, following and including an index ED admission, for future ED visits and inpatient care, were estimated to be higher for individuals residing in care homes compared to those living in the community, except for care home residents who were under 65 years of age or who had no comorbid conditions. Frequent ED visits from long-term care or care home residents have been reported previously, and often these occurred for the same reason [18]. Frequent re-attendance at ED will be partly due to increased healthcare needs due to frailty and multi-morbidities but might also be a result of healthcare needs not being addressed adequately in ED settings for this vulnerable group of patients.

NHS Greater Glasgow and Clyde is the largest health board area in Scotland, generally believed to be a good representation of the population in Scotland, however we do acknowledge that care should be taken when generalising our findings to the entire Scottish population. We were only able to control for observed confounding, which was limited to the information available from our linked data. The absence of detailed clinical information and care home characteristics can lead to residual confounding, which we could not include in our analyses. Administrative, electronic health records pose risks of information bias due to mistakes in coding and missing data. Careful data cleaning and pre-processing were undertaken to mitigate these risks. For our analysis we had to infer care home residency from an indicator in the PIS data as this is not routinely recorded in other routine healthcare data. Individuals residing in care homes who had unscheduled hospital visits were included using the first mention of a care home flag from PIS to indicate care home residency, without consideration of changes to this code (only about 5% of people entering care homes return to independent living [19]). People residing in care homes with no community prescribing records would not be included as care home residents using this approach, however we believe this to be a very small proportion of people.

Better integration of health and social care (including associated electronic records) would improve access to information that could usefully inform any interventions to improve patient care. In addition, patterns of ED visits need to be better understood in order to identify those attendances that could be managed outside ED settings and to better target interventions. This includes information on timing of ED visits to evaluate if this coincides with weekends or times during which there might be fewer staff to respond to care home staff concerns, as previous work has shown the majority occur out of hours [20].

## CONCLUSIONS

Nursing or care home residents or people staying in residential care are susceptible to unexpected alterations in their health, either because of chronic illnesses or new health events. Any decisions concerning ED attendance and potential hospitalisation must consider and balance patient and carer preferences and the medical risk. Our results indicate a notable increase in healthcare expenditure for individuals residing in care homes compared to those living in the community following unplanned acute care incidents, emphasising the importance of developing interventions that are specifically designed to meet the unique requirements of this demographic, including the use of more digital tools, as recently shown for NHS England [21].

## Data Availability

Individual-level datasets can be requested for use in other research studies through an application to NHS West of Scotland Safe Haven (https://www.researchdata.scot/safe-haven-services)

## ACKNOWLEDGEMENTS

We thank all the patients in NHS Greater Glasgow and Clyde. We are grateful for the support provided by West of Scotland Safe Haven in providing the study dataset. A secure data storage and analysis environment was provided by the Robertson Centre for Biostatistics, School of Health and Wellbeing, University of Glasgow. Thank you also to Jenni Burton and Beatrix von Wissmann for their insights into the Scottish care home landscape and care for the elderly.

## RIGHTS RETENTION STATEMENT

For the purpose of open access, the author(s) has applied a Creative Commons Attribution (CC BY) licence to any Author Accepted Manuscript version arising from this submission.

## AUTHOR CONTRIBUTIONS

Paper conceptualisation and writing: Claudia Geue

Data analysis: Govardhan Reddy Mukka, Giorgio Ciminata

Project supervision: Claudia Geue, Daniel Mackay, Giorgio Ciminata

Obtained funding for oGRE: Sandosh Padmanabhan

oGRE conceptualization: Sandosh Padmanabhan, Jill Pell

Statistical oversight and steering group: Jim Lewsey, Daniel Mackay, Ruth Dundas, Sandosh Padmanabhan

oGRE data organisation and wrangling leads: Jocelyn Friday, Tran QB Tran

oGRE data science and statistical analysis: Jocelyn Friday, Tran QB Tran, Denise Brown, Fred Ho, Claire Hastie, Mike Fleming, Claudia Geue, Govardhan Reddy Mukka, Giorgio Ciminata

Secure data storage and analysis environment management and coordination: Alan Stevenson oGRE management and coordination: Clea du Toit

## SUPPLEMENTARY MATERIALS

**Table S1:** Regression output - two-part model total costs (ED and inpatient)

|  | Probability |  | Cost ratios |  |
| --- | --- | --- | --- | --- |
|  | (first modelling part) |  | (second modelling part) |  |
|  | Coefficient (95% CI) | SE | Coefficient (95% CI) | SE |
| Cohort |  |  |  |  |
| From community | Reference |  |  |  |
| From care home | -0.226 (-0.272 to -0.181) | 0.023 | 0.006 (-0.042 to 0.053) | 0.024 |
| Year (cost incurred) | -0.213 (-0.223 to -0.203) | 0.005 | 0.003 (-0.010 to 0.017) | 0.007 |
| Sex |  |  |  |  |
| Female | Reference |  |  |  |
| Male | 0.015 (-0.028 to 0.058) | 0.022 | -0.008 (-0.055 to 0.039) | 0.024 |
| Age Category (years) |  |  |  |  |
| ≤50 | Reference |  |  |  |
| 51-55 | -0.212 (-0.435 to 0.011) | 0.114 | -0.239 (-0.514 to 0.035) | 0.140 |
| 56-60 | -0.138 (-0.346 to 0.071) | 0.106 | 0.100 (-0.269 to 0.469) | 0.188 |
| 61-65 | -0.132 (-0.337 to 0.074) | 0.105 | -0.057 (-0.336 to 0.223) | 0.143 |
| 66-70 | 0.013 (-0.183 to 0.209) | 0.100 | 0.105 (-0.142 to 0.352) | 0.126 |
| 71-75 | -0.036 (-0.210 to 0.139) | 0.089 | -0.275 (-0.498 to -0.052) | 0.114 |
| 76-80 | -0.053 (-0.208 to 0.102) | 0.079 | -0.013 (-0.268 to 0.242) | 0.130 |
| 81-85 | -0.104 (-0.257 to 0.048) | 0.078 | 0.334 (0.089 to 0.580) | 0.125 |
| 86-90 | -0.030 (-0.189 to 0.129) | 0.081 | 0.227 (0.027 to 0.428) | 0.102 |
| >90 | -0.141 (-0.321 to 0.039) | 0.092 | 0.557 (0.325 to 0.789) | 0.119 |
| Comorbidities |  |  |  |  |
| 0 | Reference |  |  |  |
| 1 | 0.765 (0.595 to 0.935) | 0.087 | 0.500 (0.265 to 0.734) | 0.120 |
| >1 | 1.292 (1.112 to 1.472) | 0.092 | 1.132 (0.919 to 1.346) | 0.109 |
| Deprivation Quintile (SIMD) |  |  |  |  |
| 1 – most deprived | Reference |  |  |  |
| 2 | -0.011 (-0.071 to 0.048) | 0.031 | 0.047 (-0.019 to 0.112) | 0.034 |
| 3 | -0.122 (-0.183 to -0.061) | 0.031 | -0.017 (-0.081 to 0.046) | 0.033 |
| 4 | -0.116 (-0.182 to -0.050) | 0.034 | 0.026 (-0.046 to 0.098) | 0.037 |
| 5 – least deprived | -0.181 (-0.247 to -0.115) | 0.034 | -0.105 (-0.180 to -0.030) | 0.038 |
| Died during follow-up |  |  |  |  |
| Alive | Reference |  |  |  |
| Dead | 1.042 (0.645 to 1.439) | 0.203 | 0.780 (0.471 to 1.090) | 0.158 |
| Interaction: age-mortality |  |  |  |  |
| ≤50 | Reference |  |  |  |
| 51-55 | -0.259 (-0.864 to 0.346) | 0.309 | 0.064 (-0.488 to 0.616) | 0.282 |
| 56-60 | -0.187 (-0.701 to 0.326) | 0.262 | -0.296 (-0.752 to 0.160) | 0.233 |
| 61-65 | -0.449 (-0.923 to 0.025) | 0.242 | -0.282 (-0.680 to 0.116) | 0.203 |
| 66-70 | -0.715 (-1.153 to -0.278) | 0.223 | -0.299 (-0.663 to 0.066) | 0.186 |
| 71-75 | -0.639 (-1.059 to -0.219) | 0.214 | -0.222 (-0.563 to 0.119) | 0.174 |
| 76-80 | -0.712 (-1.123 to -0.301) | 0.210 | -0.263 (-0.598 to 0.071) | 0.171 |
| 81-85 | -0.774 (-1.181 to -0.367) | 0.208 | -0.570 (-0.894 to -0.246) | 0.165 |
| 86-90 | -0.807 (-1.215 to -0.399) | 0.208 | -0.469 (-0.793 to -0.145) | 0.165 |
| >90 | -0.884 (-1.300 to -0.468) | 0.212 | -0.586 (-0.927 to -0.245) | 0.174 |
| Interaction: age-comorbidities (1) |  |  |  |  |
| ≤50 | Reference |  |  |  |
| 51-55 | -0.129 (-0.476 to 0.218) | 0.177 | 0.183 (-0.242 to 0.608) | 0.217 |
| 56-60 | -0.506 (-0.853 to -0.159) | 0.177 | 0.157 (-0.408 to 0.723) | 0.289 |
| 61-65 | -0.481 (-0.798 to -0.163) | 0.162 | 0.046 (-0.423 to 0.514) | 0.239 |
| 66-70 | -0.404 (-0.698 to -0.111) | 0.150 | -0.081 (-0.440 to 0.279) | 0.184 |
| 71-75 | -0.397 (-0.655 to -0.139) | 0.132 | 0.233 (-0.100 to 0.566) | 0.170 |
| 76-80 | -0.567 (-0.802 to -0.332) | 0.120 | -0.017 (-0.366 to 0.332) | 0.178 |
| 81-85 | -0.493 (-0.719 to -0.267) | 0.115 | -0.217 (-0.539 to 0.104) | 0.164 |
| 86-90 | -0.594 (-0.824 to -0.364) | 0.117 | -0.138 (-0.429 to 0.153) | 0.149 |
| >90 | -0.605 (-0.838 to -0.372) | 0.119 | -0.293 (-0.608 to 0.021) | 0.160 |
| Interaction: age-comorbidities (>1) |  |  |  |  |
| =<50 | Reference |  |  |  |
| 51-55 | -0.150 (-0.534 to 0.234) | 0.196 | 0.163 (-0.254 to 0.581) | 0.213 |
| 56-60 | -0.256 (-0.575 to 0.063) | 0.163 | -0.125 (-0.556 to 0.305) | 0.219 |
| 61-65 | -0.523 (-0.832 to -0.214) | 0.158 | -0.121 (-0.478 to 0.236) | 0.182 |
| 66-70 | -0.535 (-0.814 to -0.257) | 0.142 | -0.327 (-0.660 to 0.006) | 0.170 |
| 71-75 | -0.578 (-0.829 to -0.326) | 0.128 | -0.078 (-0.370 to 0.215) | 0.149 |
| 76-80 | -0.497 (-0.729 to -0.266) | 0.118 | -0.322 (-0.651 to 0.007) | 0.168 |
| 81-85 | -0.340 (-0.569 to -0.112) | 0.117 | -0.545 (-0.840 to -0.249) | 0.151 |
| 86-90 | -0.447 (-0.678 to -0.216) | 0.118 | -0.418 (-0.680 to -0.155) | 0.134 |
| >90 | -0.353 (-0.591 to -0.114) | 0.122 | -0.689 (-0.965 to -0.414) | 0.141 |

**Table S2:** Annual inpatient cost estimates per patient: overall and marginal by covariate using recycled predictions

|  | Community-dweller<br>£ (95% CI) | Care home resident<br>£ (95% CI) |
| --- | --- | --- |
| **Overall cost** |  |  |
| Unadjusted | 3,464 (3,334 to 3,593) | 3,861 (3,690 to 4,032) |
| Adjusted | 3,907 (3,769 to 4,044) | 3,332 (3,189 to 3,475) |
| **Sex** |  |  |
| Female | 3,344 (3,211 to 3,478) | 4,118 (3,915 to 4,322) |
| Male | 3,297 (3,139 to 3,455) | 4,074 (3,860 to 4,287) |
| **Age Category (years)** |  |  |
| =<50 | 5,579 (4,457 to 6,701) | 8,234 (6,122 to 10,346) |
| 51-55 | 4,780 (3,550 to 6,010) | 6,873 (4,674 to 9,073) |
| 56-60 | 4,962 (3,831 to 6,092) | 6,619 (4,675 to 8,563) |
| 61-65 | 3,844 (3,240 to 4,448) | 4,936 (3,956 to 5,916) |
| 66-70 | 3,833 (3,378 to 4,287) | 4,669 (4,045 to 5,292) |
| 71-75 | 3,399 (3,100 to 3,698) | 4,326 (3,882 to 4,770) |
| 76-80 | 3,374 (3,137 to 3,611) | 4,127 (3,771 to 4,484) |
| 81-85 | 3,332 (3,115 to 3,549) | 3,657 (3,413 to 3,901) |
| 86-90 | 3,451 (3,239 to 3,664) | 3,899 (3,641 to 4,156) |
| >90 | 3,446 (3,134 to 3,757) | 3,638 (3,335 to 3,940) |
| **Comorbidities** |  |  |
| 0 | 1,386 (1,252 to 1,520) | 1,437 (1,282 to 1,591) |
| 1 | 2,705 (2,529 to 2,881) | 2,920 (2,707 to 3,133) |
| More than 1 | 4,855 (4,646 to 5,063) | 5,449 (5,196 to 5,702) |
| **Deprivation Quintile (SIMD)** |  |  |
| 1 – most deprived | 3,355 (3,208 to 3,502) | 4,149 (3,938 to 4,360) |
| 2 | 3,568 (3,320 to 3,815) | 4,415 (4,072 to 4,757) |
| 3 | 3,210 (2,984 to 3,436) | 3,964 (3,674 to 4,253) |
| 4 | 3,409 (3,143 to 3,674) | 4,213 (3,862 to 4,564) |
| 5 – least deprived | 2,859 (2,621 to 3,097) | 3,520 (3,211 to 3,829) |

**Table S3:** Annual ED cost estimates per patient: overall and marginal by covariate using recycled predictions

|  | **Community-dwellers**<br>**£ (95% CI)** | **Care home resident**<br>**£ (95% CI)** |
| --- | --- | --- |
| **Overall cost** |  |  |
| Unadjusted | 799 (774 to 824) | 1,130 (1,084 to 1,177) |
| Adjusted | 928 (898 to 958) | 944 (913 to 975) |
| **Sex** |  |  |
| Female | 802 (774 to 830) | 1,121 (1,077 to 1,166) |
| Male | 806 (775 to 837) | 1,130 (1,082 to 1,179) |
| **Age Category (years)** |  |  |
| =<50 | 2,167 (1,818 to 2,517) | 2,928 (2,354 to 3,502) |
| 51-55 | 1,462 (1,128 to 1,795) | 2,036 (1,508 to 2,563) |
| 56-60 | 1,055 (901 to 1,210) | 1,447 (1,180 to 1,715) |
| 61-65 | 813 (709 to 918) | 1,106 (923 to 1,289) |
| 66-70 | 802 (729 to 876) | 1,053 (940 to 1,166) |
| 71-75 | 730 (683 to 777) | 966 (894 to 1,038) |
| 76-80 | 775 (725 to 825) | 1,017 (953 to 1,081) |
| 81-85 | 756 (719 to 793) | 941 (895 to 988) |
| 86-90 | 781 (739 to 823) | 972 (919 to 1,025) |
| >90 | 798 (726 to 869) | 929 (869 to 990) |
| **Comorbidities** |  |  |
| 0 | 455 (432 to 477) | 572 (539 to 605) |
| 1 | 729 (693 to 765) | 917 (871 to 963) |
| More than 1 | 1,196 (1,130 to 1,261) | 1,504 (1,413 to 1,596) |
| **Deprivation Quintile (SIMD)** |  |  |
| 1 – most deprived | 863 (831 to 896) | 1,205 (1,157 to 1,253) |
| 2 | 825 (782 to 868) | 1,152 (1,088 to 1,216) |
| 3 | 720 (676 to 765) | 1,022 (958 to 1,085) |
| 4 | 745 (699 to 792) | 1,053 (984 to 1,123) |
| 5 – least deprived | 673 (631 to 716) | 954 (891 to 1,017) |

